# Death in People with Down syndrome: Mortality statistics and novel predictors in US Medicaid and Medicare enrolled adults

**DOI:** 10.64898/2026.07.17.26358090

**Authors:** Salina Tewolde, Anthony J Rosellini, Amy Michals, Brian G. Skotko, Juan Fortea, Bernard Khor, Samuel Handelman, Eric Rubenstein

## Abstract

People with Down syndrome have higher age-specific mortality rates compared to the general population as well as peers with other intellectual and developmental disabilities. While a large proportion of mortality is attributable to Alzheimer’s disease, many die prior to Alzheimer’s diagnosis and some live to old ages, dying without Alzheimer’s. Our objectives were to use 11 years of Medicaid and Medicare data to describe characteristics and factors related to death in adults with Down syndrome and use machine learning to identify which conditions most strongly predict death in the full population and stratified by age. We identified death using Center for Medicare and Medicaid Systems reported date of death health conditions using ICD 9 and 10 codes. We used a case-control design with risk set sampling to have that controls to mimic the distribution of times of incident Alzheimer’s disease. We trained gradient boosted trees to identify strongest predictors. Our cohort included 137,293 adults with Down syndrome. Among those, 30,894 (22.5%) died during the study period. Mean age at death among those who died was 55 years (SD=10). Mean age of death in those with Alzheimer’s disease was 59 (SD=7) and those without was 52 (SD=12). The most influential predictors of mortality were any claim for dementia, any claim for pneumonia, re-occurring claim for cardiovascular disease three years before index death, and any claim for heart failure and epilepsy. Our results align with previous clinical work and highlight intervenable areas to reduce mortality in the Down syndrome population.

## Introduction

Down syndrome (DS) is a genetic condition caused by the full or partial triplication of chromosome 21 (1). In the US, there are over 100,000 adults in living with Down syndrome (2). Individuals with DS have shorter life expectancies compared to the general population, but survival has extended from an average life expectancy of 5 years in the 1950’s to 60 years in 2020 (3). Advances in management of congenital heart defects, medical technology, and overall population health have increased survival rates in this population (1). Nevertheless, people with DS have higher age-specific mortality rates compared to the general population as well as peers with other intellectual and developmental disabilities (4).

A key cause of premature mortality in adults with DS is Alzheimer’s disease (Down syndrome Alzheimer’s Disease [DS-AD]). DS and DS-AD are closely linked through the triplication of chromosome 21, where the increased genetic dosage of the APP gene cause early-onset Alzheimer’s disease (5). By age 40, nearly all individuals with DS show neuropathological changes consistent with Alzheimer’s Disease and the lifetime risk for developing symptoms of dementia is more than 90% (5). Furthermore, the onset of Alzheimer’s disease in this population occurs 20-30 years earlier than the general population, contributing to greater morbidity and mortality (6). While prevalent, DS-AD is only one component of multi-faceted causal mechanisms that lead to death in Down syndrome. In this instance, DS-AD as a chronic condition may increase risk of death from other acute conditions, such as pneumonia (7).

Many adults with DS die prior to expected DS-AD diagnosis and some live to old ages, dying without DS-AD, illustrating that mortality in this population is not solely driven by DS-AD (8). Understanding what other pathways contribute to premature death in this DS population is crucial for improving early clinical diagnosis and management of co-occurring conditions that may prevent early death. People with DS develop more chronic health conditions compared to those without DS (9). Studies have shown that those with DS have higher prevalence of epilepsy, hearing loss, and hematologic and oncologic disorders than in disomic individuals (9). While these risk factors and co-occurring conditions are well documented in this population, less is known about the timing and onset of how these conditions contribute to mortality risk.

Previous studies have found associations between mortality and certain comorbidities in adults with DS that vary by age and dementia status. Most existing studies looking at causes of death in DS focus on individual co-occurring conditions and lack a sufficient cohort size (10), (11), (12). Death certificate data are flawed, with most common cause of death among people with Down syndrome being DS, not capturing cause of death and do not characterize co-occurring conditions surrounding death (13).

Machine learning methods have gained increasing attention for their ability to improve disease prediction and diagnosis (14). By leveraging administrative datasets, supervised machine learning approaches can model complex nonlinear relationships and interactions in their predictions that may not be easily captured in traditional statistical models (14). One potential application of supervised machine learning is to utilize health insurance data from the majority of the DS population in the US that receive benefits through Medicaid and Medicare. In the US, the majority of adults with DS rely on public health insurance for their medical care. Our objectives for this study were to use 11 years of Medicaid and Medicare data in the US to (1) describe characteristics and factors related to death in adults with DS and (2) use machine learning to identify which conditions most strongly predict death in both the full population with DS and stratified by those younger and older than 40 years.

## Methods

### Data source

We used data from the Down Syndrome Toward Optimal Trajectories and Health Equity using Medicaid Analytic eXtract (DS-TO-THE-MAX) project. DS-TO-THE-MAX is a longitudinal administrative claims cohort following all adults ≥18 years of age with DS enrolled in Medicaid and/or Medicare at any point between 2011 and 2022. Datafiles included inpatient, outpatient-, and long-term care files for Medicaid and Medicare. DS was identified by examining claims from any source for ICD-9 code 758.0 or ICD-10 codes Q90.0, Q90.1, Q90.2, and Q90.9. More details on cohort derivation are presented in Rubenstein et al. (15), (16). This project was considered exempted research by the local Institutional Review Board.

### Inclusion and exclusion criteria

We included all adult enrollees in Medicaid and/or Medicare who were enrolled for at least one year between 2011-2022 and met the criteria for DS identification. Our criteria meant we excluded individuals who died in their first year of claims or had a death date previous to their first year of data.

### Outcome identification

Death was identified using the beneficiary date of death recorded in Medicare and Medicaid administrative enrollment files. For those who did not die during the study period, a random time point was assigned as an index date to serve as a reference point for identifying predictors (i.e. incidence density sampling) (17).

### Co-occurring conditions

We used International Classification of Disease-9 and 10 diagnostic codes and procedure codes to identify 104 chronic and acute health conditions operationalized as binary predictors. To identify common and chronic conditions, we use established algorithms from the Chronic Conditions Data Warehouse (CCW) (18), (19) (eTable1). To capture timing of when conditions occurred relevant to death or index date, we coded conditions as ever occurring, within three months, six months, one year, two years, and three years prior to death or index date. We labeled variables to be incident to capture new diagnoses and reoccurring for conditions that could have more than one claim (e.g., pneumonia). To be incident or recurrent, an individual would need to have a condition-specific period of time without claims for the condition. Conditions were also grouped together into broader categories (cancer, mental health, cardiovascular disease, bone break) (Supplement 1). Demographic variables were excluded from the model to avoid mixing clinical conditions with administrative data. After pre-processing the data and removing rare conditions (those that had less than 10 deaths per condition), our final model was processed using 104 unique conditions comprising 610 variables.

### Demographic variables

Demographic variables were from the demographic enrollment files in Medicaid and the Master Beneficiary Summary File in Medicare. We aligned data from Medicaid and Medicare, so all data was consistent regardless of what program one was enrolled in. When race/ethnicity was missing in Medicaid (∼25%) we used multiple imputation to impute race/ethnicity (See Rubenstein et al. 2024 for more details [16]). We created binary indicators for each racial/ethnic group, sex, region, health insurer (Medicaid, Medicare, or Dual enrolled), and age category at index date in ten-year increments.

### Statistical and Machine Learning analysis

We first calculated descriptive statistics for demographic variables among our total cohort and then stratified by those who died and did not die within our study period. We constructed a lifetable and Kaplan Meier curve documenting the probability of death in five-year intervals. We then calculated univariate odds ratios (ORs) via logistic regression for all conditions and demographic variables to understand the association between predictors and death.

We trained and tested an extreme gradient boosting (XGBoost) model to predict mortality using all conditions and demographic variables. XGBoost was used due to its ability to handle large datasets (e.g., computational efficiency) and capture nonlinear relationships and interactions (20), (21) and was implemented using the *caret* package in R (22). The dataset was randomly split into training (80% n=109,834) and testing (20% =27,459) subsets. Within the training set, the model hyperparameters (e.g., tree depth, node size) were tuned using 10-fold cross validation grid search. The optimal model was then evaluated in the test dataset. We calculated sensitivity, specificity, and receiver operating curve (AUC) to identify the most optimal model. To evaluate whether predictions varied by age, we ran the same model for those 40 years and younger and those over 40.

We calculated SHapley Additive exPlanations (SHAP) values using the *shapviz* package to interpret the model and assess the contribution and interactions of top predictors for mortality (23). SHAP is a model-agnostic method for interpreting machine learning models by estimating each feature, or predictors, contribution. Each SHAP value describes how a predictor influences mortality risk or protection. A higher mean absolute SHAP value indicates that the variable was more influential in predicting mortality. We used Beeswarm plots and univariate ORs to determine the directionality of predictive associations.

We evaluated the necessity of using a flexible tree-based learner by comparing to flexible and traditional linear-based approaches including elastic net regression and logistic regression. We compared models by evaluating AUCs and visualized distributions using boxplots.

## Results

Our cohort included 137,293 adults with DS enrolled in Medicaid and/or Medicare at any time from 2011-2022. Among those, 30,894 (22.5%) died during the study period (Table 1). Mean age at study entry of those who died was 50 years (SD=10) and 31 years (SD=13) among those who did not die. Mean age at death among those who died was 55 years (SD= 10) and mean age at index date for those that did not die was 35 years (SD=14). Among those who died, 52.7% were male, 83.3% were non-Hispanic white, and 90.5% were enrolled in both Medicaid and Medicare (Table 1). Of those who didn’t die, 48.8% were male, 70.8% were non-Hispanic white, and 40.4% were enrolled in both Medicaid and Medicare (Table 1).

**Table 1:**
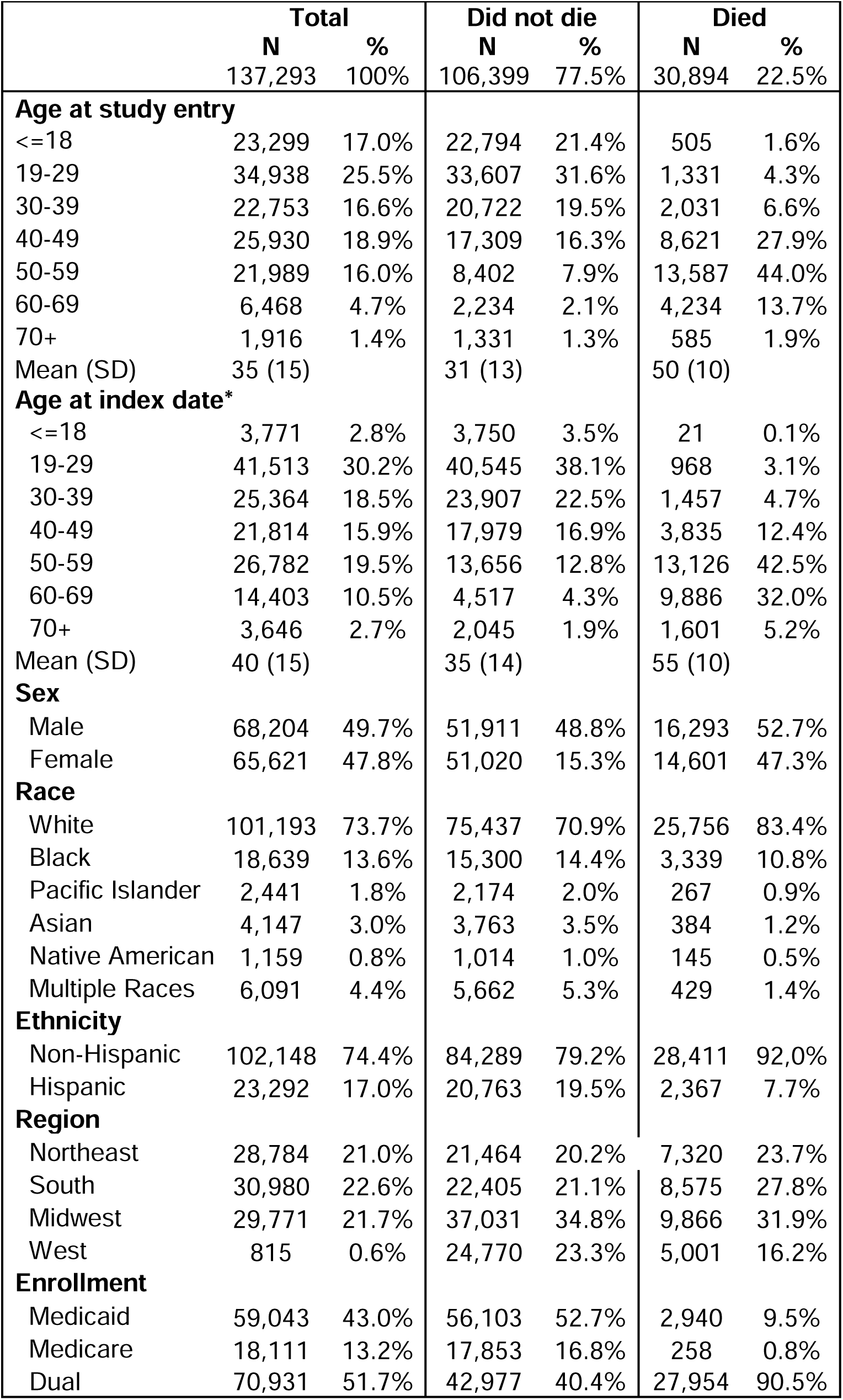
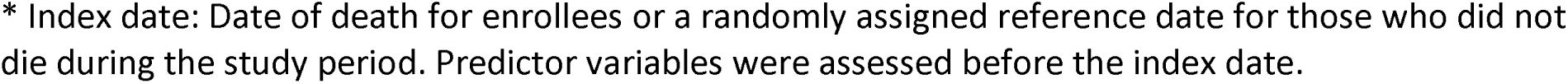
Demographic characteristics of adults with Down syndrome enrolled in Medicaid and/or Medicare, 2011-2022.

|  | Total |  | Did not die |  | Died |  |
| --- | --- | --- | --- | --- | --- | --- |
|  | N | % | N | % | N | % |
|  | 137,293 | 100% | 106,399 | 77.5% | 30,894 | 22.5% |
| **Age at study entry** |  |  |  |  |  |  |
| <=18 | 23,299 | 17.0% | 22,794 | 21.4% | 505 | 1.6% |
| 19-29 | 34,938 | 25.5% | 33,607 | 31.6% | 1,331 | 4.3% |
| 30-39 | 22,753 | 16.6% | 20,722 | 19.5% | 2,031 | 6.6% |
| 40-49 | 25,930 | 18.9% | 17,309 | 16.3% | 8,621 | 27.9% |
| 50-59 | 21,989 | 16.0% | 8,402 | 7.9% | 13,587 | 44.0% |
| 60-69 | 6,468 | 4.7% | 2,234 | 2.1% | 4,234 | 13.7% |
| 70+ | 1,916 | 1.4% | 1,331 | 1.3% | 585 | 1.9% |
| Mean (SD) | 35 (15) |  | 31 (13) |  | 50 (10) |  |
| **Age at index date\*** |  |  |  |  |  |  |
| <=18 | 3,771 | 2.8% | 3,750 | 3.5% | 21 | 0.1% |
| 19-29 | 41,513 | 30.2% | 40,545 | 38.1% | 968 | 3.1% |
| 30-39 | 25,364 | 18.5% | 23,907 | 22.5% | 1,457 | 4.7% |
| 40-49 | 21,814 | 15.9% | 17,979 | 16.9% | 3,835 | 12.4% |
| 50-59 | 26,782 | 19.5% | 13,656 | 12.8% | 13,126 | 42.5% |
| 60-69 | 14,403 | 10.5% | 4,517 | 4.3% | 9,886 | 32.0% |
| 70+ | 3,646 | 2.7% | 2,045 | 1.9% | 1,601 | 5.2% |
| Mean (SD) | 40 (15) |  | 35 (14) |  | 55 (10) |  |
| **Sex** |  |  |  |  |  |  |
| Male | 68,204 | 49.7% | 51,911 | 48.8% | 16,293 | 52.7% |
| Female | 65,621 | 47.8% | 51,020 | 15.3% | 14,601 | 47.3% |
| **Race** |  |  |  |  |  |  |
| White | 101,193 | 73.7% | 75,437 | 70.9% | 25,756 | 83.4% |
| Black | 18,639 | 13.6% | 15,300 | 14.4% | 3,339 | 10.8% |
| Pacific Islander | 2,441 | 1.8% | 2,174 | 2.0% | 267 | 0.9% |
| Asian | 4,147 | 3.0% | 3,763 | 3.5% | 384 | 1.2% |
| Native American | 1,159 | 0.8% | 1,014 | 1.0% | 145 | 0.5% |
| Multiple Races | 6,091 | 4.4% | 5,662 | 5.3% | 429 | 1.4% |
| **Ethnicity** |  |  |  |  |  |  |
| Non-Hispanic | 102,148 | 74.4% | 84,289 | 79.2% | 28,411 | 92.0% |
| Hispanic | 23,292 | 17.0% | 20,763 | 19.5% | 2,367 | 7.7% |
| **Region** |  |  |  |  |  |  |
| Northeast | 28,784 | 21.0% | 21,464 | 20.2% | 7,320 | 23.7% |
| South | 30,980 | 22.6% | 22,405 | 21.1% | 8,575 | 27.8% |
| Midwest | 29,771 | 21.7% | 37,031 | 34.8% | 9,866 | 31.9% |
| West | 815 | 0.6% | 24,770 | 23.3% | 5,001 | 16.2% |
| **Enrollment** |  |  |  |  |  |  |
| Medicaid | 59,043 | 43.0% | 56,103 | 52.7% | 2,940 | 9.5% |
| Medicare | 18,111 | 13.2% | 17,853 | 16.8% | 258 | 0.8% |
| Dual | 70,931 | 51.7% | 42,977 | 40.4% | 27,954 | 90.5% |

\* Index date: Date of death for enrollees or a randomly assigned reference date for those who did not die during the study period. Predictor variables were assessed before the index date.

The Kaplan Meier curve illustrating survival in our cohort is presented in Figure 1B. The median survival time was approximately 60 years with an interquartile range of 57 years to 67 years. When assessing age of death by DSAD status (Figure 1A) mean age of death in those with AD was 59 (SD=7) and those without was 52 (SD=12).

**Figure 1:**
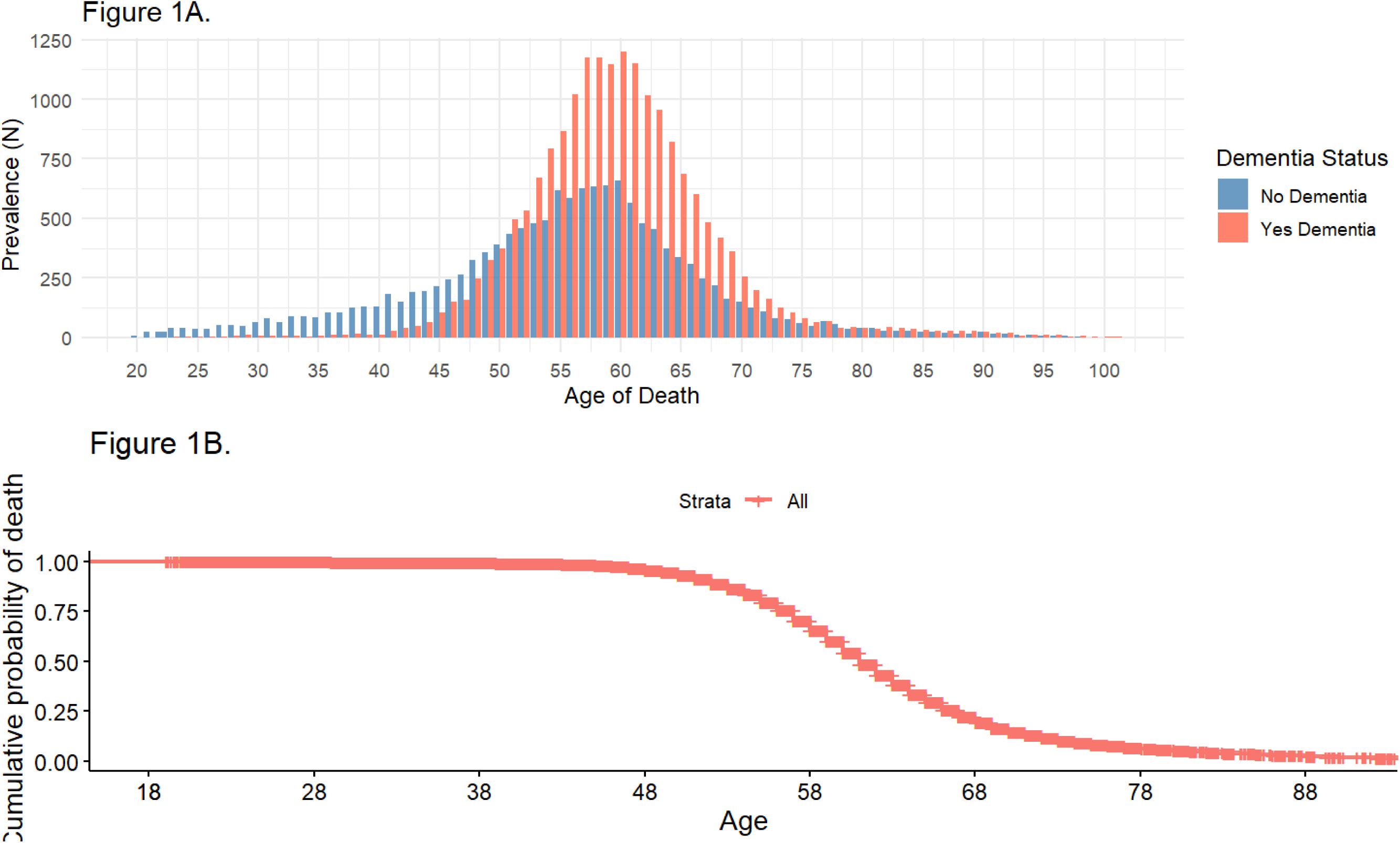
Age of death for adults with Down syndrome enrolled in Medicaid and/or Medicare, 2011-2022. Figure 1A: Age of death by dementia status Figure 1B: Kaplan–Meier Survival Curve for Age at Death

In unadjusted odds ratios, conditions with the largest odds for death were for any claim of DS-AD (OR: 17.1, 95% CI: 16.5, 17.6), any claim of ulcer (OR: 15.0, 95% CI: 14.2, 15.8), and any claim of pneumonia (OR: 8.4, 95% CI: 8.1, 8.6) (eTable 2). The top protective factor was being overweight two-years pre-index date (OR: 0.09, 95% CI: 0.06, 0.12).

After hyperparameter tuning and model training, the final XGBoost model achieved an AUC of 0.93 CI: (0.92-0.94) in the test sample. Individuals in the highest quintile of predicted risk had an observed mortality rate of 88% corresponding to a positive predictive value of 88% (eFigure1). Based on the SHAP values, the most influential predictors of mortality were any claim for dementia, any claim for pneumonia, re-occurring claim for cardiovascular disease three years before index death, and any claim for heart failure and epilepsy (Figure 2). Among the top predictors, a history of recurrent cardiovascular disease was associated with lower predicted mortality in the model (SHAP values >0: increase predicted risk, SHAP values <0 decreases predicted risk) (Figure 2). To evaluate model performance, we compared our XGBoost model to elastic net and logistic regression models. XGBoost achieved the highest AUC in the test sample (0.94) followed by elastic net (0.93), and logistic regression (0.86) (eFigure2).

**Figure 2:**
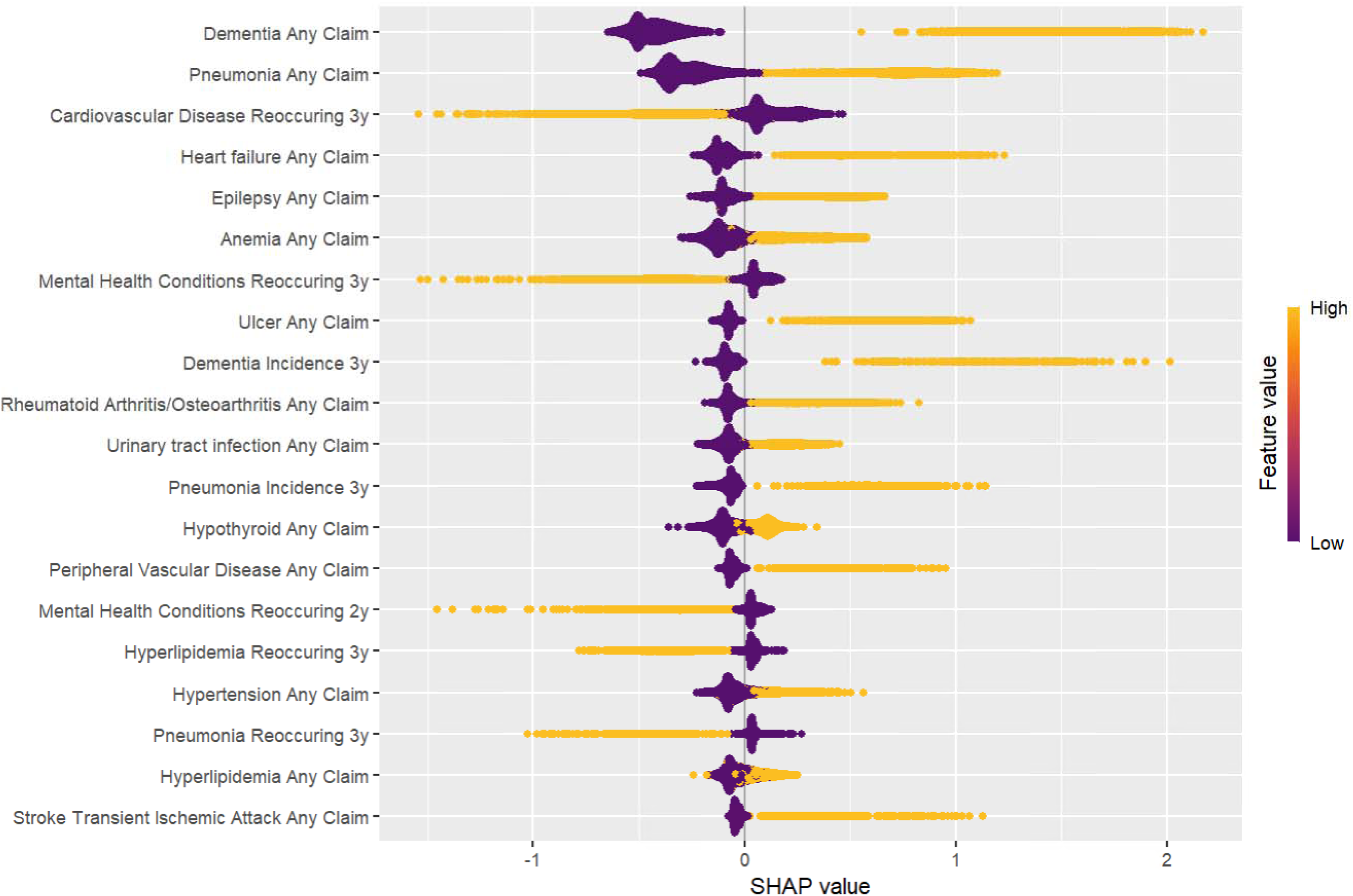
Beeswarm plot for SHAP values of 20 most influential variables predicting death among adults with Down syndrome enrolled in Medicaid and/or Medicare. SHAP value plots for each feature are shown, ranked by importance (mean absolute SHAP value). Each dot represents 1 individual, with their position representing their SHAP value. Color is used to indicate the specified feature characteristic. Yellow dots indicate those who have the listed characteristic, while purple dots indicate those who do not. For example, for the any claims for pneumonia, individuals with a yellow dot have claims for pneumonia and appear predominantly to the right of Y-axis, indicating higher predicted risk for mortality, while those with a purple dot do not have claims for pneumonia.

To assess if predictors varied by age, we re-trained the model stratified to those <40 years and those ==40 years. For the under 40 group, the top predictors were having any claim for pneumonia, re-occurring claim for cardiovascular disease three years before index death, re-occurring claim for mental health conditions three years before index death, and having any claim for heart failure and anemia (Figure 3). A history of recurrent cardiovascular disease and mental health claims were associated with lower predicted mortality. When looking at those 40 and older, having any claim for dementia, pneumonia, epilepsy, re-occurring claim for hyperlipidemia three years before index death, and having any claim for ulcers contributed to increased predicted risk of mortality (Figure 3). Hyperlipidemia three years before index death was protective and associated with lower predicted mortality whereas the other top predictors were associated with a higher predicted mortality.

**Figure 3.**
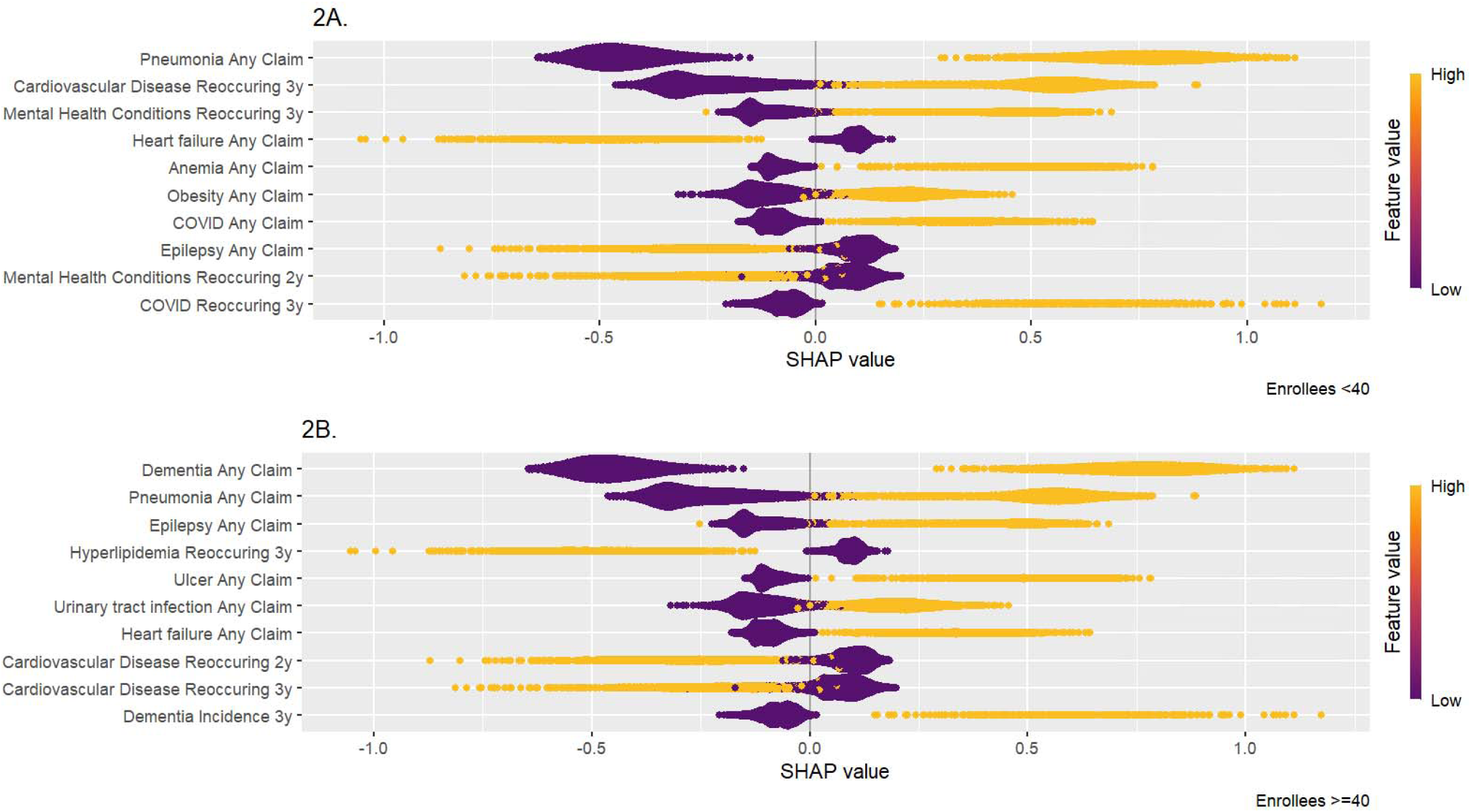
Beeswarm plot predicting death among adults with Down syndrome enrolled in Medicaid and/or Medicare stratified by age in years. SHAP beeswarm plots showing the most influential predictors of mortality among adults with Down syndrome stratified by age group. Each dot represents 1 individual, with their position representing their SHAP value. Color is used to indicate the feature value. Yellow dots indicate those who have the condition (high feature value), while purple dots indicate those who do not (low feature value). SHAP values represent the contribution of each predictor to mortality risk, where positive values indicate increased predicted risk of death and negative values indicate decreased risk. Among adults aged ≥40 years, dementia, pneumonia, and epilepsy were among the strongest predictors of mortality. Among adults aged <40 years, pneumonia, recurrent cardiovascular disease, and mental health conditions were the strongest predictors

## Discussion

Adults with DS die earlier than peers with death largely attributed to DSAD (24). However, death with AD is often a combination of multiple factors and many die without DSAD. We used a large claims data set to describe death in adults with DS and use machine learning to identify most impactful factors that predict mortality. Findings have implications for individual clinical care and population health approaches for adults with DS.

Mean age of death in our data aligns with most recent estimates in the Down syndrome population of 59 years, although we do not have data on death in childhood which may drive down our average (25), (26). There were few deaths before 50 years of age, highlighting a relatively resilient period in mid-life with low risk of mortality before AD onset. Previous work has examined racial and ethnic disparities in DS mortality, finding little differences in adulthood.

Many of the strongest predictors of death are aligned with our current knowledge about DS (11). The AD connection is well established, as is the increased mortality risk due to infectious inflammatory diseases, such as pneumonia. Heart failure is likely associated with congenital heart defects and epilepsy often occurs surrounding AD diagnosis (27). Urinary tract infection, ulcers, and arthritis have immunological facets that may especially harm people with DS (28). We hypothesize that protective factors such as mental health conditions were associated with younger age, and cardiovascular conditions with more health care use. This finding is in line with machine learning models predicting death in the general population, which did not find mental health conditions to be strong predictors of mortality in population data (29).

Our findings have implications for healthcare. Identifying when conditions begin to contribute to mortality risk may help delay or prevent early death in people with DS. For example, early detection and management of epilepsy, may help prevent complications that contribute to early mortality (30) (31). The mortality risk of pneumonia is greatly reduced by vaccination; key opportunities in DS include combating under-utilization of vaccines and rationally managing their advanced immune aging with earlier adoption of strategies reserved for older typical individuals (32).

## Limitations

Claims data reflects healthcare utilization and administrative billing practices that may not fully capture all underlying medical conditions. Although we used Medicare validated algorithms to identify conditions, there still may be misclassification. Additionally, index date was defined as date of diagnosis rather than actual onset of disease, which might introduce reverse causation. To mitigate potential time bias, we created windows of observation for all conditions observed. Our analysis was also limited to lack of laboratory test results, longitudinal clinical measurements, and other phenotypic data that could provide additional context. Risk set sampling was used to account for differing person time and loss to follow up. Residual bias may still exist due to survival-individuals with poorer health may have died earlier and had less time for conditions to be documented.

## Conclusion

Healthcare claims data are an advancement to understanding cause of death in DS compared to flawed death certificates data. Identifying and intervening on conditions that lead to death, among people with and without DS-AD, may help us prolong the lifespan for all people with DS and support continued preventive care and routine health screening before dementia onset.

## Supporting information

Supplement 1, eFigure 1+2

## Data Availability

Data are accessed under a data use agreement with the Centers for Medicare and Medicaid services and are not avaialble for reuse.

https://www.resdac.org

