## Supplement 1, eFigure 1+2 for "Death in People with Down syndrome: Mortality statistics and novel predictors in US Medicaid and Medicare enrolled adults"

Supplement 1: Condition groupings

1. Mental Health: Depression, Attention-deficit/hyperactivity disorder (ADHD), Schizophrenia, Post-traumatic stress disorder (PTSD), Depression, Bipolar disorder, Anxiety disorder
2. Cardiovascular Disease (CVD): Hyperlipidemia, Heart failure, Hypertension, Ischemic heart disease, Stroke, Peripheral vascular disease
3. Cancer: Leukemia, Breast cancer, Colorectal cancer, Prostate cancer, Lung cancer, Any cancer
4. Bone Break: Ankle fracture, Wrist fracture, Tibia/fibula fracture, Femur fracture, Hip fracture, Shoulder fracture, Vertebral fracture, Hip or pelvic fracture

Supplement eFigure1: Positive predictive value of Mortality Risk

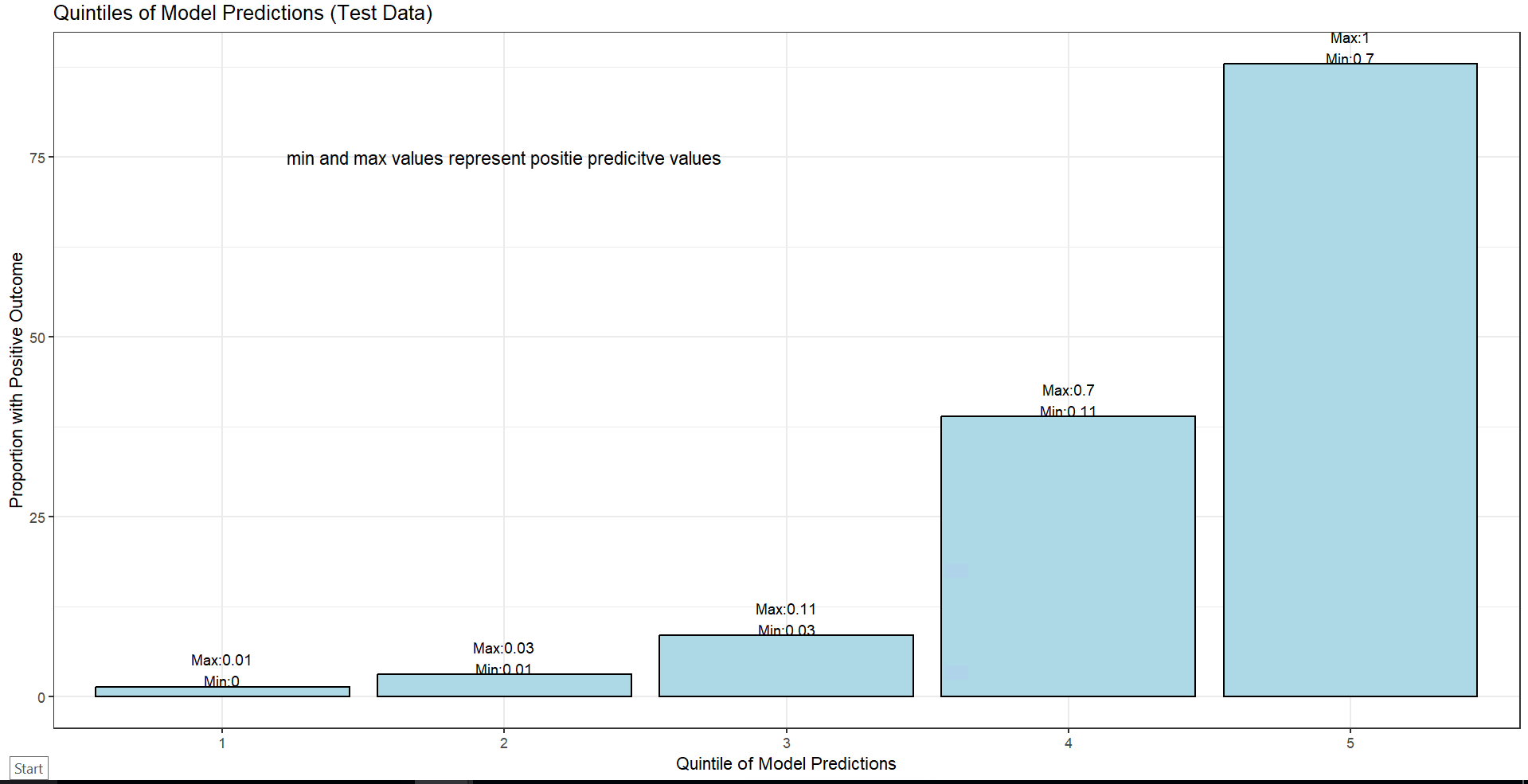

Supplement eFigure2: Model Comparison (XGBoost, Logistic Regression, ElasticNet)

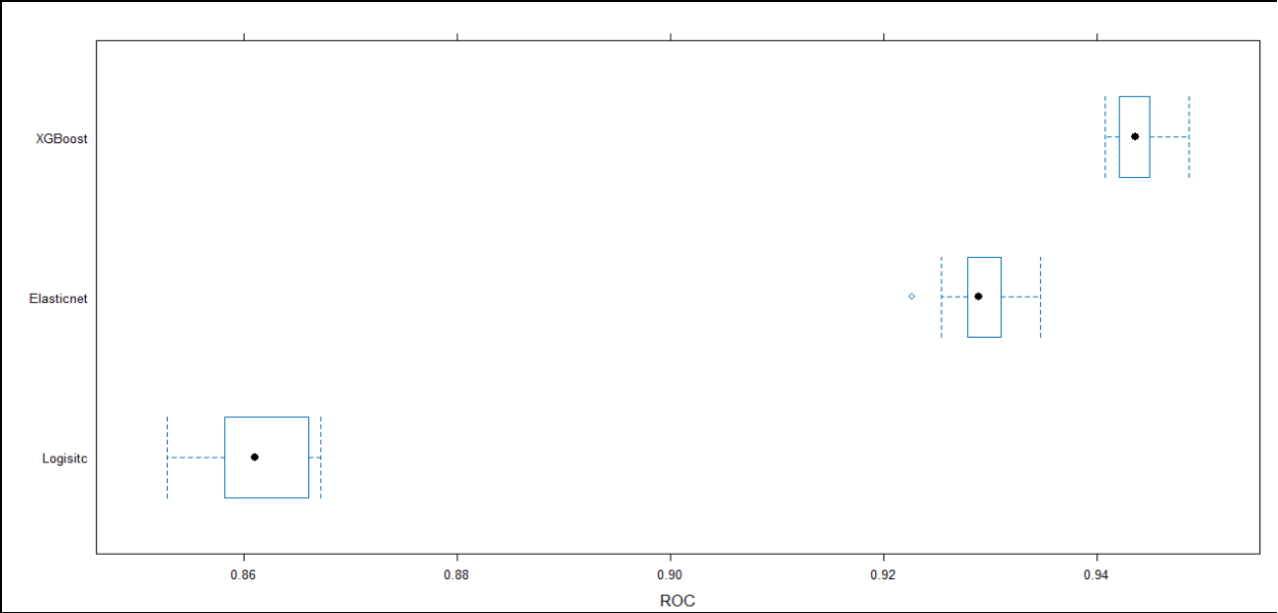
